# Leveraging Open-Source Solutions to Build a Low-Cost Digital Pathology Pipeline for Translational Research

**DOI:** 10.64898/2026.04.25.26350240

**Authors:** Joseph Stenberg, Aparna Gullapalli, Kathryn Foucar, Daniel Babu, Jordan Redemann, Nancy Joste, Charles Foucar, Dita Gratzinger, Tracy George, Robert Ohgami, Rama R. Gullapalli

## Abstract

Digital Pathology (DP) is a fast-emerging branch of pathology focused on digitizing pathology data. A key challenge of DP usage for pathology laboratories, especially mid- to small-sized clinical labs, are the upfront costs associated with instrumentation and the logistical challenges of implementation. In the current project, we built an end-to-end DP solution using low-cost, open-source components that is user-friendly at a small scale. We repurposed readily available microscopy components in a pathology lab to assemble a fully functional DP pipeline for translational research applications. We tested multiple low-cost complementary metal-oxide semiconductor (CMOS) cameras in this project and chose a user-friendly Canon camera for image acquisition. An open-source DP server solution, OMERO v.5.6.4, was used as the image management system (IMS) to host and serve the WSIs on an Ubuntu 22.04 operating system. The server-hosted WSI images were evaluated remotely and asynchronously by multiple pathologists physically situated in Albuquerque, NM; Salt Lake City, UT; and Palo Alto, CA. Each pathologist assessed the quality of the WSI pipeline, image quality, and WSI interaction experience using a 23-question survey. Overall, the custom, low-cost WSI pipeline was noted to be a robust and user-friendly experience by the pathologists. The current DP setup is unlikely to be useful as a commercial, scalable DP pipeline for large-scale clinical applications. However, it demonstrates the feasibility of creating customized, small-scale DP solutions (at a low price point) for asynchronous translational pathology research applications. Additionally, building customized DP pipelines provides excellent educational opportunities for pathology residents to gain in-depth knowledge of the various technical elements of a DP workflow. In summary, we have established a low-cost, end-to-end WSI DP pipeline useful for spatiotemporally asynchronous translational pathology research, in an academic setting.

## I. Introduction

Digital pathology (DP) represents an ongoing revolution in pathology practice, a transformative extension of traditional glass-slide microscopy that pathologists are comfortable with. Advances in histological slide scanning technologies and computational capacities allow greater integration of DP platforms into routine clinical practice. Whole-slide imaging (WSI), a keystone element of DP technology, creates high-resolution, digital versions of conventional histologic slides. Ongoing improvements in WSI scanner performance, data processing algorithms, and rapidly declining image storage costs are facilitating increasing adoption of digital pathology workflows into clinical practice.

The digital format of WSIs extends routine histopathological evaluation by a pathologist into additional forms of pathology review, such as remote/robotic telepathology consultation, increased inter-institutional collaborations, and advanced imaging analytics to gain novel information content from patient slides. Artificial intelligence (AI)- driven analytics are further supercharging these transformations in pathology ^1–3^. The clinical advantages of an automated DP workflow are numerous. Online WSI images enable immediate, 24 x 7 remote access for pathologists, alleviating challenges associated with physical slide delivery and the need for a pathologist’s physical presence. DP assisted workflows can help to address the current shortages of pathologist expertise in the US and across the globe ^4–7^.

AI-assisted annotation tools on DP platforms can enable precise labeling, such as highlighting and measuring specific areas of interest (e.g., tumor size, distance to inked margins, and foci of lymphovascular invasion), which play a key role in patient healthcare. All of this DP data can then be made readily available to external consultants and clinical colleagues to enable better diagnostic outcomes for patients. It has been shown that rapid digital access to WSI cases can reduce turnaround times for external consultations and enable earlier therapeutic interventions ^8^. Pathologists can lessen their dependence on microscopes and finalize cases off-site, supporting a more flexible practice with potential cost-saving measures 8. When properly implemented, these benefits can greatly improve patient-centered care while making the clinical practice more efficient.

WSI has faced much skepticism over the years for technical issues pertaining to its image quality and diagnostic reliability when compared to the gold standard of light microscopy. However, the computational capabilities of digital pathology slide scanners have evolved enormously over the past decade and continue to rapidly improve each year. There are many studies in the literature reporting diagnostic concordance rates of 90-95% as compared to the light microscope^9–11^. The College of American Pathologists (CAP) has actively supported the use of WSI for primary diagnosis, further reinforcing the push towards clinical adoption ^12^. Digitization of glass slides transforms static tissue sections into analyzable datasets, permitting integration with the orthogonal methods of genomic, proteomic, and clinical datasets of a patient. Furthermore, all of this data is readily assessable for advanced AI-driven analytics, which can provide deeper insights into a patient’s healthcare. Over the past decade, AI-based, deep learning models have been shown to be increasingly capable of tumor detection, automated grading, and histologic subtype classification tasks, all directly from routine hematoxylin and eosin-stained sections ^1,13–16^. More recently, advanced AI/deep learning models have demonstrated the ability to predict underlying molecular alterations, including mutational status, limited gene expression signatures, and prognostic outcomes from routine histomorphological slides ^13,17–20^. A rapidly developing area of research in pathology involves the use of spatial analytic models to assess spatial variation in tissue tumor burden, degree of immune cell infiltration, and spatial tumor microenvironment architecture patterns ^21–23^. Together, these advances illustrate that WSI has surpassed the diagnostic equivalence with traditional microscopy by demonstrating non-inferior diagnostic concordance with light microscopy alone and serving as a novel, emerging platform for AI-driven pathologic analysis^7,8,24^.

A further advantage of DP workflows is the availability of novel avenues for interinstitutional research collaborations and medical education in pathology^5,25–28^. For example, remote image hosting and browser-based reviews can facilitate departmental collaborations across institutions worldwide, thereby eliminating barriers to knowledge sharing^5,27^. In addition, DP workflows deliver greater flexibility for annotation, asynchronous review, and better accessibility to share AI models across different institutions ^24^. Similar benefits are also widely observed through the use of DP in medical/pathology education efforts. During the COVID-19 pandemic, medical education programs rapidly transitioned to remote and/or hybrid models to maintain continuity of pathology training while prioritizing safety^29^. Although pandemic effects have largely subsided and are a distant memory now, many educational institutions have expanded the use of DP workflows for medical education activities ^25,29,30^. Within pathology residencies, academic centers have implemented virtual microscopy for sign-out sessions, allowing greater flexibility for slide annotation, simultaneous slide driving between attending and resident, and an overall greater convenience in returning to prior cases for educational reviews ^31^. Traditional slides are no longer used in the American Board of Pathology examinations, thereby requiring trainees to have a baseline competency in elements of virtual microscopy. In parallel with the ongoing shift toward digitization in pathology, there is an increasing need for pathology trainees to be familiar with learning, adopting, and translating diagnostic and technical skills between WSI workflows and traditional microscopy workflows^5,6,8^.

The major stumbling block to the widespread clinical adoption of DP are the pricing costs, available technical skills (pathology informatics), and regulatory obstacles^32^. While leading academic centers (with adequate financial resources) have implemented hybrid and/or fully digital workflows, smaller pathology practices are unable to make these transitions due to financial and technical challenges. Some key challenges include high upfront capital expenditures for WSI scanners, personnel training, and software integration into existing information technology infrastructures ^33,34^. Other highlighted concerns include diagnostic variability with specific specimens, regulatory compliance, and patient data security issues ^24,35^. Although most studies show DP workflows can improve efficiency, some have reported negative impacts ^36^.. Many of these concerns pertaining to DP workflows in a clinical setting are heightened by ongoing budgetary constraints and overall declining healthcare reimbursement. The substantial costs and infrastructure requirements of many commercial WSI systems limit access for many small laboratories, creating significant disparities and barriers to widespread DP adoption in clinical healthcare.

In the current project, our objective is to develop, implement, test, and evaluate a cost-effective, manual WSI pipeline which can be used by small-scale laboratories and/or research groups. In the software space, a viable alternative to commercial software solutions are the open-source solutions. Since the inception of the internet, the open-source community has provided highly robust alternatives to closed, costly commercial software products. Some such examples are – 1. Adobe (commercial) vs Gimp (open-source), 2. Microsoft WORD (commercial) vs LibreOffice (open-source) and 3. EndNote (commercial) vs Zotero (open-source). We strongly feel that similar open-source solutions must exist in the space of digital pathology to increase affordability (and increase experimentation) for interested individuals/departments. The current project summarizes our efforts in the use of readily available hardware and software components to build one such DP pipeline. Our DP workflow system incorporates a manual region-of-interest (ROI) acquisition and stage movement procedure for individual image registration following by WSI creation. The overarching goal is to explore a practical, reproducible model for DP workflows in a setting where purchases of high-throughput scanners is not financially viable. The current study does not aim to replace high-throughput commercial scanners for primary clinical DP, but rather to explore a pragmatic, research-oriented DP workflow alternatives that are amenable to learning and experimentation. To support the stated goals of accessibility and flexible design, we have leveraged open-source software solutions and image management systems for modular scalability without vendor lock-in at each step of the process. We anticipate this project to serve as a starting-point and a reproducible blueprint for other pathology translational research labs to build DP workflows suited for their own specific research needs, while simultaneously offering educational value to pathology trainees in understanding the key elements of digital pathology.

## II. Materials and Methods

### i. Whole Slide Imaging (WSI) Platform – Computational Hardware

The open-source DP slide-imaging system was assembled using an in-house bright-field microscope, coupled to a DSLR (Digital Single-Lens Reflex; Canon; Model EOS 6D) camera. Prior to settling on the Canon 6D, we tested the image acquisition protocols on multiple CMOS cameras – i. Basler acA-1940-40GC (Sony IMX249 sensor), ii. PointGrey GS3-PGE-23S6C-C (Sony IMX174 sensor) and iii. Pixelink D7512CU (Sony IMX253 sensor). See Supplemental Figure 1 for additional information on the cameras. All of the cameras and microscope components were acquired as used equipment, primarily through secondary-market vendors (eBay), or were already available in the Gullapalli lab. The microscope used in this study is a routine Olympus BX41 instrument with an add-on trinocular head. While the microscope had multiple objectives (4X, 10X, 20X, 40X, and 60X), we primarily used an Olympus UPlanSApo 10×/0.40 objective for image acquisition in this study. The Canon EOS-6D full-frame camera attached to the microscope using an Olympus U-SPT microscope phototube adaptor with an internal relay lens (Olympus NFK 3.3X LD). A custom assembled desktop computer was built using components purchased from multiple vendors (see table 1). The key computer configuration includes - an Intel Core i9 CPU, 32GB of DDR4 RAM, and an NVIDIA GeForce GTX 1660 Super graphics card. Full configuration details are provided in Table 1.

**Table 1:** A cost breakdown of the various computational and microscopic components used to build the custom, open-source WSI slide imaging system.

| Item | Component Details | Price/Source |
| --- | --- | --- |
| **Computer** | Intel® Core™ i9-9820X CPU @ 3.30GHz | \$120/eBay |
| | 32GB DDR4 RAM | \$60/eBay |
| | Nvidia GeForce GTX 1660 Super | \$105/eBay |
| | Gigabyte X299 UD4 Pro-CF MB | \$100/eBay |
| | 1 TB NVMe Internal SSD | \$60/eBay |
| | 650-Watt Power Supply Unit | \$60/Newegg |
| | ATX Mid-Tower PC Case | \$75/eBay |
| **Microscope** | Olympus BX41 Microscope with Trinocular head, 4X, 10X, 20X, 40X and 60X Objectives | Department Property |
| | Olympus UPlanSApo 10×/0.40 Objective (Optional) | \$1,000/eBay |
| **Camera** | Cannon EOS-6D Full Frame Camera | \$350/eBay |
| | Fotasy Adjustable T2 Lens to Cannon EF EFS Adapter | \$10/Amazon |
| | Fotasy 35-90mm M42 to M42 Lens Macro Helicoid Adapter | \$30/Amazon |
| | Relay Lens | \$40/eBay |
| | Olympus U-SPT Microscope Phototube Camera Adapter | \$100/eBay |
| **Software** | Canon EOS Utility Software | Free/Canon |
|  | OMERO – Open Microscopy Environment | Open-source GPL |
|  | FIJI – Image Stitching Software | Open-source GPL |
|  | Custom stitching software – Python | Open-source GPL |

### ii. Whole Slide Imaging Platform – Computational Software

Images were captured using the Canon EOS Utility Software using a tungsten white balance preset, an exposure (shutter speed) of 1/10 second, and an ISO of 320. The Köhler illumination was enabled on the microscope at the start of each imaging session. A 3 × 3 mm region of interest was photographed in a serpentine acquisition pattern (left-to-right, right-to-left) with approximately 20-30% overlap between adjacent fields. Images from each case were stitched into a single file using the open-source Fiji image processing package 21. A visual representation of the photographing and imaging stitching process is shown in Figure 1A&B. The nine stitched whole slide images were blinded and stored on a Ubuntu 22.04 LTS Linux server, which also served as the central repository for the remote slide images. The server was configured as a headless node managed via the open-source image management software (IMS) platform OMERO (version 5.6.0). A 4TB network-attached storage (NAS) device was configured to store the backup WSIs. The OMERO web client (version 5.29.1) enabled secure remote slide review across participating institutions. A graphical representation of the slide imaging pipeline is shown in Figure 2.

**Figure 1:**
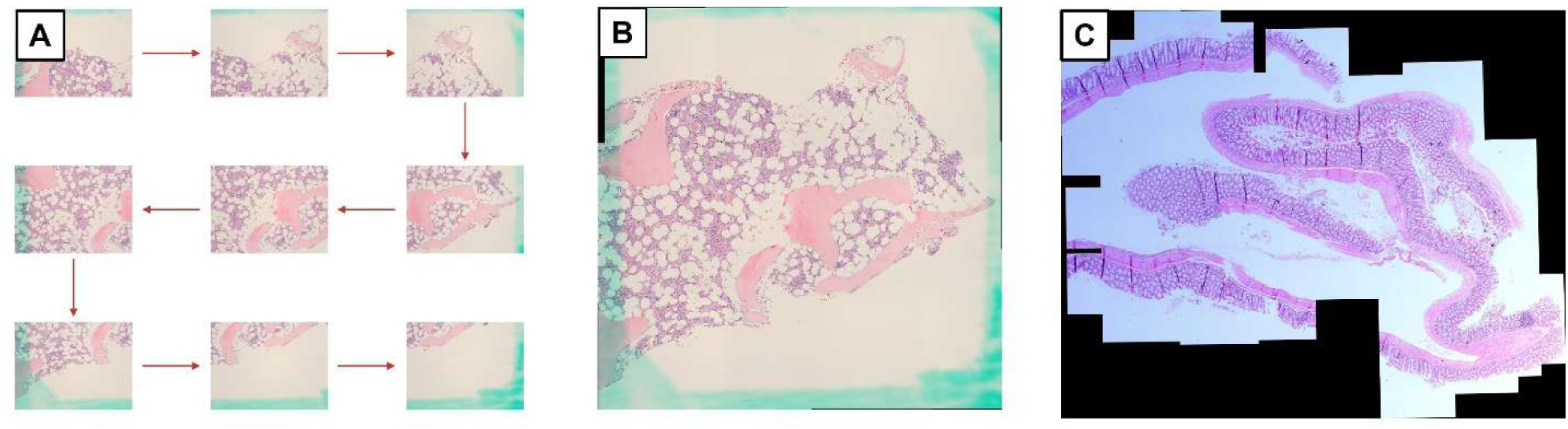
The custom WSI workflow process developed in this project using open-source, readily available components. Image stitching was performed using the Fiji software. H and E slides are imaged initially, followed by image collection on the microscope set up with approximately 20-30% overlap between successive images. The stitched images are managed by an OMERO Image Management System (IMS) located on an Ubuntu 22.04 LTS operating server, which can be accessed remotely by various end-user pathologists.

**Figure 2:**
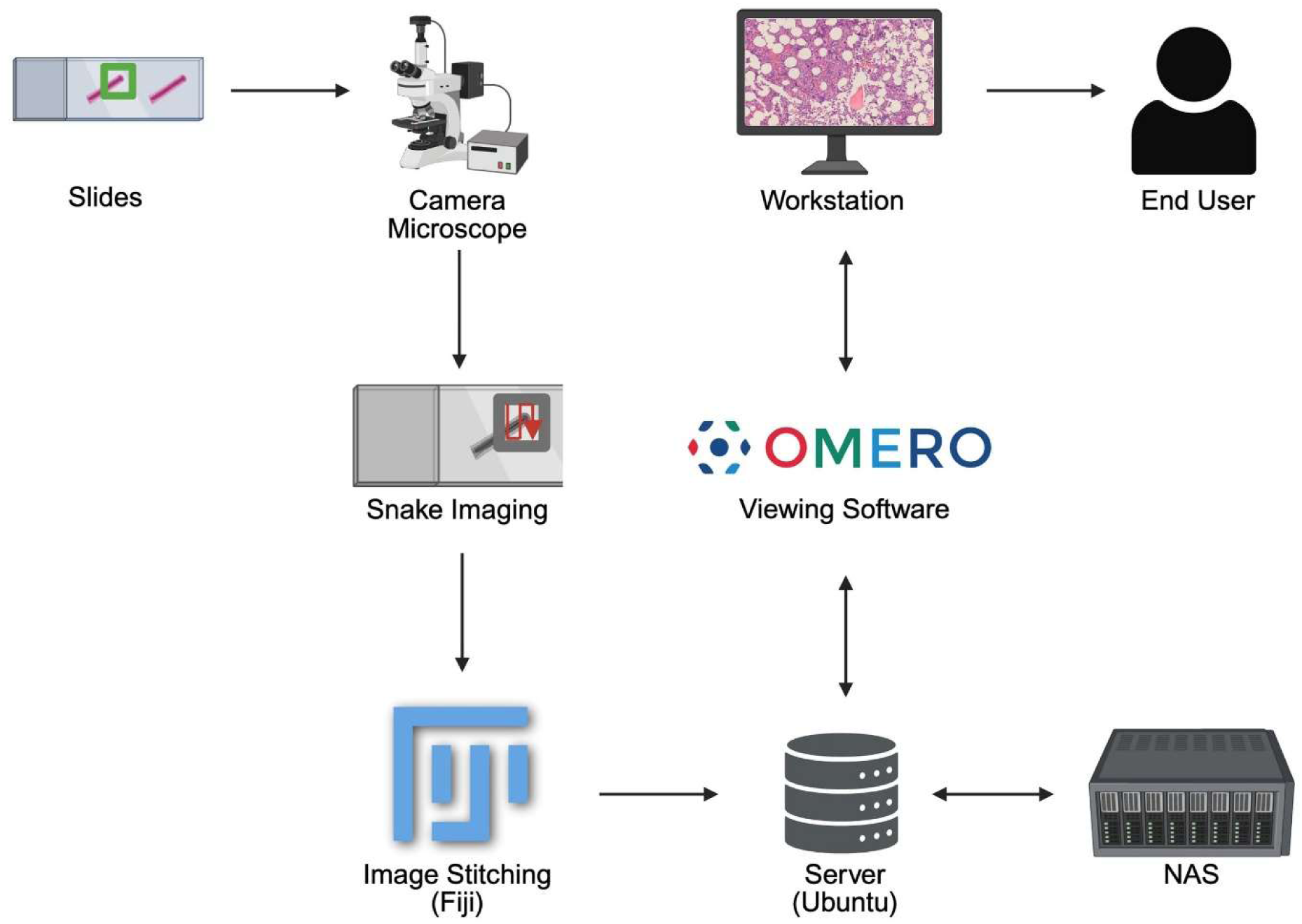
An example of the image stitching outcomes implemented in this project. A. Individual tiles of the bone marrow regions in a patient sample B. The complete stitched WSI outcome. C. A section of a murine colon with improved image acquisition settings.

### iii. WSI Test Case Selection Process

A selection of bone marrow biopsy cases from the University of New Mexico were identified using the PowerPath laboratory information system by searching for the terms “NORMOCELLULAR” and “MDS” in the year 2025. The “NORMOCELLULAR” cases were sequentially reviewed for the absence of significant abnormality until n = 4 was reached. The “MDS” cases were sequentially reviewed to identify cases of myelodysplastic syndrome (neoplasm) until n = 5 was reached (see Supplemental Figure 2). A single trephine biopsy slide from each of the nine cases was selected for manual scanning, as previously described. A 3 × 3 mm square was drawn with a marker pen over the bone marrow biopsy histological areas to image sufficient tissue for WSI image creation and further evaluation by the pathologists.

### iv. Pathologist Recruitment Evaluation and Slide Imaging Survey

The main goal of the current project was to test the usability of our custom, end-to-end WSI solution built in-house. We recruited seven pathologists from three institutions present at different geographical locations to test the WSI interface. The pathologists recruited in the study had diverse training backgrounds and years of pathology experience. Brief written instructions in the form of a i. WORD document, ii. an email summary to access the OMERO WSI interface, and iii. a link to the survey questions were provided to the study pathologists. No in-person instructions were provided. After the pathologists reviewed the nine scanned cases on the online OMERO platform, they proceeded to complete a survey questionnaire. We created a 23-question survey, which was designed to evaluate various elements of the opne-source WSI digital pathology pipeline. Survey categories included i. the ease of use, ii. comparison with traditional microscopy, iii. translational research usability, iv. utility of the set up in transitioning to clinical digital pathology use , and v. shortcomings of the platform. In addition to the survey responses, demographic and background data on pathology subspecialties, years of practice, and general thoughts regarding digital pathology were obtained. A separate survey section was created to assess the pathologists’ impressions regarding scanned slides and their ability to correctly classify the scanned WSI slides as myelodysplastic syndrome or without diagnostic abnormality (i.e., normal bone marrow cases). The questionnaire was published and made available to pathologists via Google Forms. The exact wording on the various survey questions is provided in Table 2. The survey responses were exported from Google Forms to Excel and CSV files for downstream evaluation, which was evaluated using custom R code (v.4.5.0) to create the various graphics presented in the main body of the text.

**Table 2:**
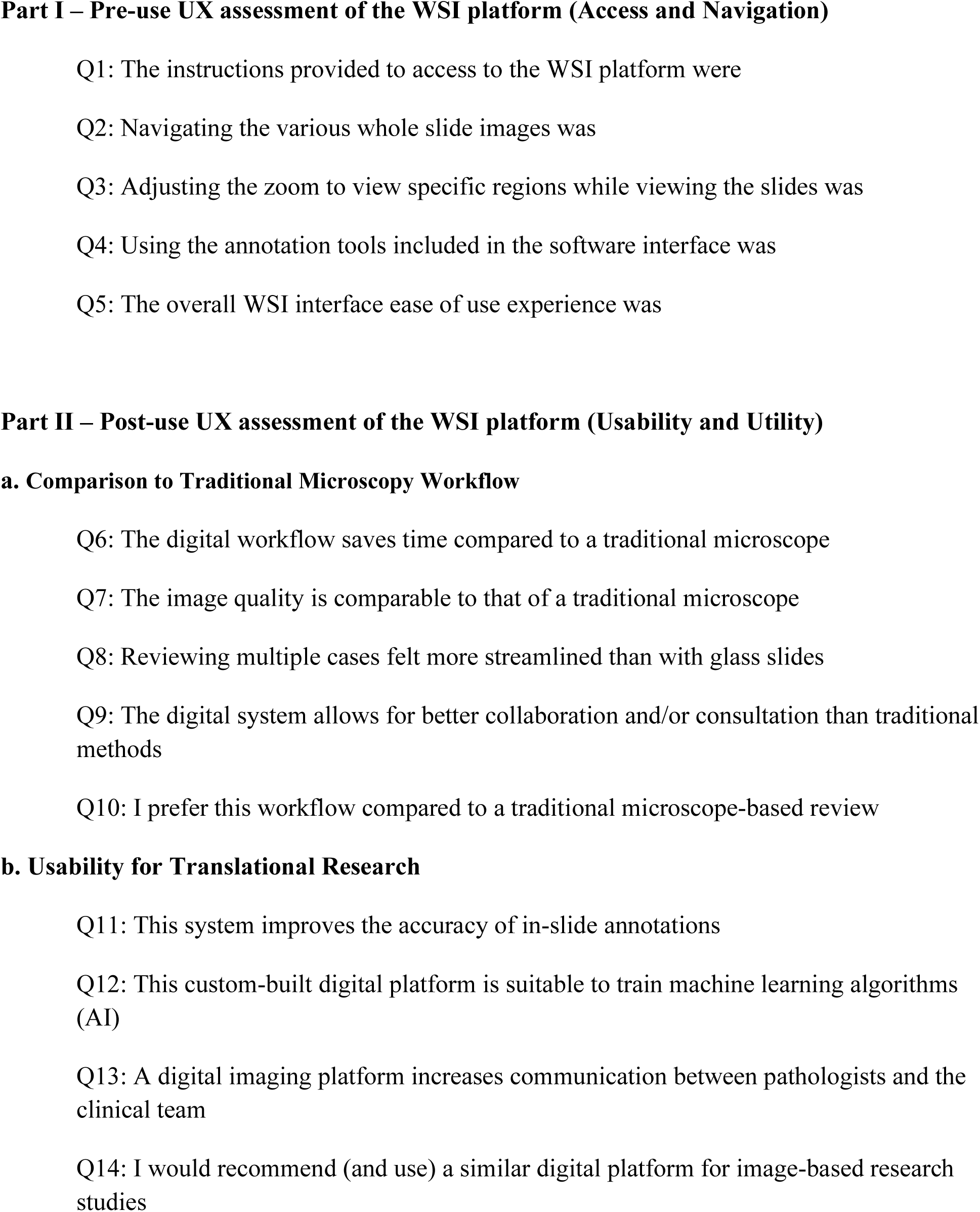
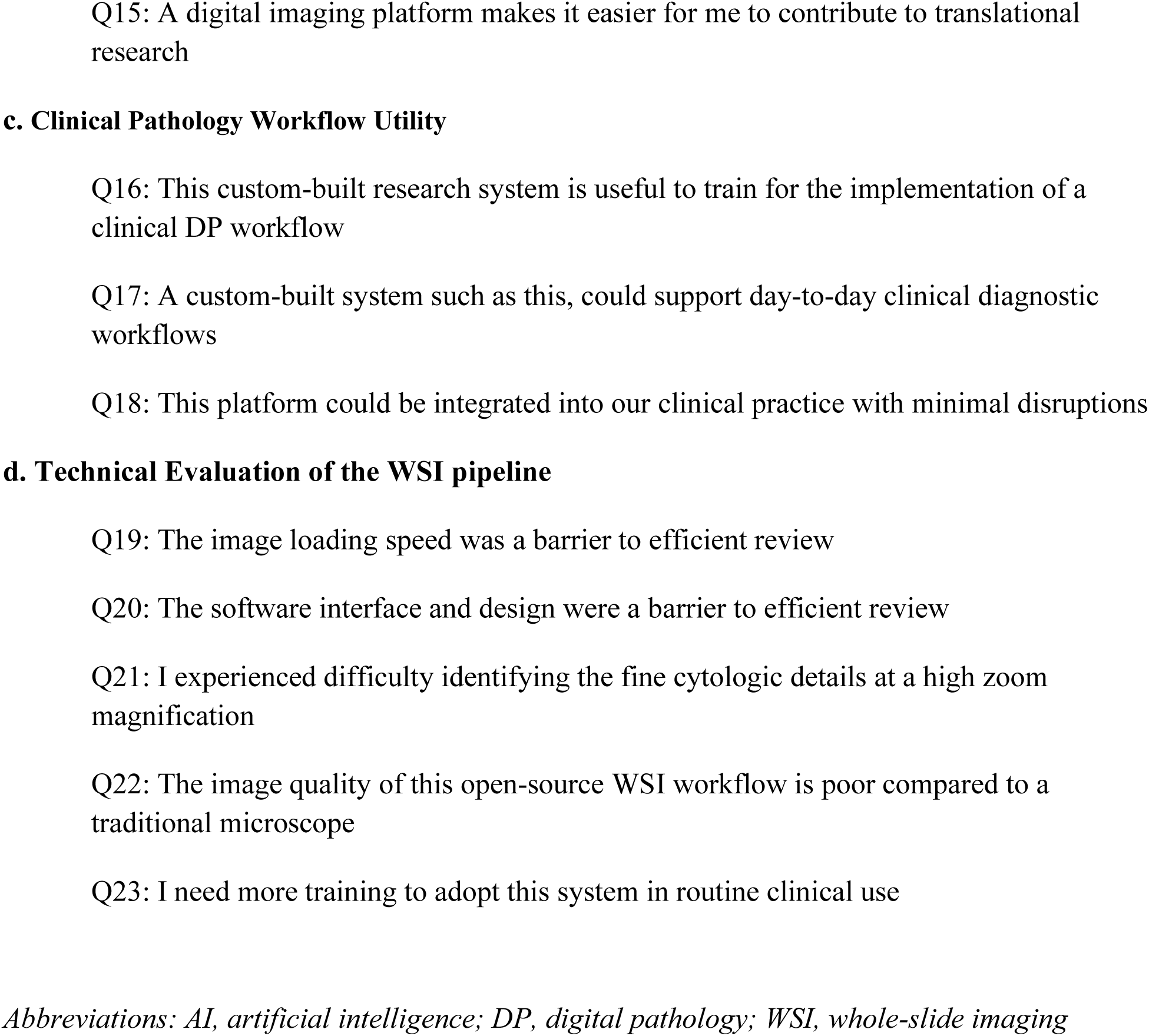
Questions used to survey the pathologists in the project. The survey focused on pre- and post-use user experience (UX) feedback collected from seven (n=7) pathologists who participated in the study. Survey questionnaire used to evaluate pathologist impressions of the custom-built digital pathology platform. All of the survey responses were evaluated using a categorical, Likert scale approach

## III. Results

### A. Whole Slide Imaging (WSI) Set-Up – Computational Hardware

A majority of the optical hardware components that were used in our digital pathology whole slide imaging platform are modular and easily available online. With the universal availability of high-end CMOS camera setups for industrial automation (and machine learning applications in automated manufacturing), there has been a revolution in the quality of the CMOS cameras and, a steep reduction in their pricing. In the current project, we initially evaluated multiple cameras available from the industrial automation market– 1. Basler (www.basler.com; Last accessed 02/10/2026) 2. Point Grey (now part of www.flir.com; Last accessed 02/10/2026) and 3. PixelLink camera (www.navitar.com; Last accessed 02/10/2026). All of these companies produce cameras routinely used in industrial automation, process control, and image evaluation purposes. These cameras were purchased from eBay and assessed for suitability in the digital pathology pipeline. The specific details of the sensor sizes, frame rates, and camera interface configuration are provided in Supplemental Figure 1. While all of the cameras generated images that were useful for whole slide imaging, the software interfaces were noted to be complicated and cumbersome for routine implementation in our setup. We then explored the possibility of using a commercial DSLR camera and chose Canon for its versatility in the hardware and software interfaces. Specifically, the Canon 6D model was chosen for the project and was noted to provide excellent quality images with fine-tuned control over the image collection process. Additionally, the Canon software interface was easy to implement with user-friendly image acquisition control. In the Canon software interface, the primary controls are the 1. image acquisition time (“shutter delay”), 2. The ISO of the image (“shutter speed”) and 3. lighting contrast. While there are many additional controls of the image acquisition setting that are native to the Canon software, we did not utilize any of them to keep the workflow simple for resident use.

The second major hardware component necessary for the WSI setup was a hardware computer capable of image acquisition via digital control. We custom-built our hardware computer primarily on the backbone of an 8-core Intel(R) Core (TM) i7-9700 CPU 3.00 GHz chip with 32 GB of RAM memory and a one terabyte hard drive for image acquisition and storage. Windows 10 was chosen to comply with the institutional software requirements and to enable enterprise oversight of the laboratory computer. Additionally, the majority of add-on software for the various hardware components used in our WSI setup works best on Windows 10, which was a key consideration. Image acquisition was via a fast USB 3.0 cable connection between the Canon 6D camera and the computer in a “live image acquisition” mode. A detailed specification of the custom-built computer hardware is provided in Table 1. The microscope used to build the WSI setup was a standard issue microscope (Olympus BX41) provided to the senior author (R.R.G). The PI subsequently upgraded multiple objectives on the microscope to include high-end UPlanSApo objectives (10X, 20X, 40X, and 60X) with high numerical apertures (10X - 0.40, 20X – 0.75, 40X – 0.95, and 60X – 0.90) to enable high-resolution imaging of superior quality. Almost 90% of the hardware necessary for the current WSI build was purchased incrementally on third party websites (i.e., eBay) at much lower prices than newer, commercially available items. It is the author’s experience (R.R.G) that almost all of the items needed to build a modular WSI setup can be acquired for a mere 10-15% of the list pricing in the form of used products. The author (R.R.G) has never had a non-useful purchase experience (for electronic items specifically) acquired from third party websites. In the rare instance that a product does not meet the specifications, eBay’s robust return policy helps protect against bad purchases.

### B. Whole Slide Imaging (WSI) Set-Up – Computational Software

The second key element required in assembling the WSI setup are the different software products and applications necessary for an end-to-end WSI setup. In our current system, we have had to evaluate software applications in three main categories – a. Image stitching software, b. Image acquisition software, and c. WSI Image Management System (IMS).

Due to the manual, asynchronous nature of the current WSI pipeline, it is critical to maintain proper data hygiene and protocols to ensure seamless utilization of the WSI pipeline. In the initial phase, we ensured that all of the slides are properly labeled with unique identifiers. The slide label specific data folders are created locally on the computer’s data hard disk with appropriate project naming conventions/specifications. All of these can vary from setup to setup based on individual preferences. However, it is critical that once the conventions are established, all members of the lab adhere to these specifications. Additional written protocols will ensure consistency in image data acquisition procedures by all members of the lab, including rotating residents/fellows.

a. Image Stitching software – The fundamental algorithmic basis for image stitching protocols has a long history of development in the computer vision community. A robust high-level description of image stitching algorithms is provided at the following link for the interested reader (see https://en.wikipedia.org/wiki/Image_stitching; Last accessed 02/10/2026). A variety of image stitching algorithms exist in the literature, such as the scale invariant feature transform (SIFT), which is an image processing algorithm to detect salient, stable feature points in an image, or the RANSAC (RANdom Sample Consensus) algorithm, which is used in creating a composite image. A detailed discussion of the algorithms is out of scope in this article. There are a large number of software applications that have implemented these algorithms to enable image stitching as a part of the overall software package. These include - Autostitch, Hugin, PTgui, Panorama Tools, Microsoft Research Image Composite Editor, CleVR Stitcher, Adobe Systems’ Photoshop, and ImageJ among others. See the following link for details on individual software applications and their algorithms. (https://en.wikipedia.org/wiki/Category:Photo_stitching_software; Last accessed 02/10/2026). In the current project, we tested and evaluated three open-source software – a. Hugin b. Microsoft Research Image Composite Editor (ICE) and c. ImageJ. While all of the software were able to provide a robust stitched final image with relative ease, we chose ImageJ for our project due to the consistent (and ongoing) development pipeline associated with this software and the widespread adoption of the ImageJ (https://imagej.net/ij/ and FIJI – https://imagej.net/software//fiji/downloads ; Last accessed 02/10/2026) software within the imaging scientist community.

b. Image Acquisition Software – The image acquisition software is determined by the eventual choice of the camera used for the platform. Since we acquired multiple cameras, we tested all of the software associated with each camera. It is worthwhile to note that industrial-grade USB cameras provide extensive, flexible options; this may be overwhelming for the novice user (such as a pathologist). In contrast, the software for a commonly used camera, such as a Canon DSLR camera, is designed with the novice user in mind, which makes the use/implementation a much easier process. While the image acquisition software specification was not our primary consideration, the ultimate choice of a Canon 6D camera provided a favorable outcome in the data collection process as an added benefit.

c. WSI Image Management System (IMS) - A key advantage of commercial WSI off-the-shelf systems is the tight-knit integration that exists between the hardware and software components in the development (and usage) process. However, this advantage is offset by the WSI system purchase costs and the recurring costs of maintenance contracts associated with such a system. A clinical system implementation (which sees heavy and constant use) usually involves planning for the annual maintenance contract costs. However, in the context of a WSI system used for research purposes (like a core facility), long-term maintenance contracts (and constant WSI system upgrades) are usually not feasible. Thus, a low-cost modular system (such as the current project) provides research users with options for flexible upgrades and avoids expensive long-term contract costs. For such users, the OMERO (Open Microscopy Environment Remote Objects; https://openmicroscopy.org/omero; Last accessed 02/10/2026) is an open-source, client-server software platform for managing, visualizing, and analyzing biological microscopy images^37,38^. OMERO is a robust, scalable solution for intensive image management such as large WSI datasets)^37–39^. We implemented the OMERO IMS as our preferred solution in our DP pipeline for translational pathology research. The server-side OMERO software was installed on an Ubuntu 22.04 LTS version Linux desktop, which hosted all of the WSI datasets used in this project. In addition to the server responsible for the upload and management of the WSI datasets, we also tested client-side access via the installation of an OMERO desktop app, which connected with the OMERO server to access the pathology images/datasets. A key feature of the OMERO software development process is client access via an interface built into a commonly used web browser, such as Firefox, Google Chrome, or Microsoft Edge. For the purposes of this project, we tested Firefox and Google Chrome interfaces, which worked perfectly to access the WSI datasets without any problem. Furthermore, we provided OMERO client access to remote pathologists in Albuquerque, NM; Palo Alto, CA; and Salt Lake City, UT.

### C. Pathologist Demographics and Pondering the Future Use of AI in Pathology

A total of seven subspecialty-trained academic pathologists were recruited for the project. The recruited pathologists had specialized sub-specialty training in hematopathology (n=6), molecular pathology (n=1), gynecologic pathology (n=1), and cytopathology (n=1). The pathologist cohort had a mean age of 53 years (range, 35-75 years) and a mean attending-level experience in pathology of ∼21 years (range, 1-47 years). In the initial part of the survey, we sought to understand the expertise level and familiarity of our pathologist cohort with issues pertaining to digital pathology and its use in clinical practice. All pathologists reported at a minimum average familiarity (57.1%) with digital pathology, and most (42.9%) reported an above-average level of familiarity with digital pathology workflows. The specific focus of this project (i.e., use of our pipeline for translational research activity) received much interest among the pathologists, with 85.8% expressing an interest in the use of digital pathology for translational research.

We then sought to evaluate the group’s thoughts on the emerging roles/use cases for digital pathology/AI in the clinical workplace over the next decade. A majority of pathologists responded by stating that by the end of the next decade, DP/AI would be important for routine pathology clinical practice, including the automation of many pathology-related tasks, such as i. quantitative immunohistochemistry (100%) ii. automated micrometastasis screening (100%), iii. primary pathology diagnosis (86%), iv. biomarker evaluation and monitoring (86%), v. automated microorganism screening (86%), vi. therapeutic selection and guidance (71%) and vii. patient outcome prognosis (71%). Overall, the pathologists indicated an important role for AI/DP in changing the way pathology will be practiced in the next decade with the automation of many of the routine tasks (e.g., IHC and micrometastasis detection) by DP/AI. Extending this application focused evaluation, we queried the group of the likelihood of pathologists being “replaced” by AI/DP, as it is one of the key concerns about AI currently, across various fields of work. The majority of pathologists felt it was somewhat unlikely (42.9%) and highly unlikely (42.9%) that DP/AI would be able to replace pathologists by the end of the next decade. We feel this optimism naturally reflects a deep understanding of the complex nature of pathology-driven diagnostic workflows and that pathologists are unlikely to be replaced by DP/AI anytime soon.

Focusing more on the technical elements of our custom, end-to-end DP workflow solution, we assessed the accuracy of pathologists in correctly identifying the disease pathology under test (MDS vs non-MDS). A total of nine bone marrow test cases WSI’s were reviewed entirely remotely via our OMERO web-based client. Of these cases, four lacked any significant histopathologic abnormality, and five had a morphological diagnosis of MDS. The overall diagnostic concordance for MDS identification on scanned slides was 84%. In eight of the nine cases, there was a consensus greater than 70%. In a single case (case # 6), only 43% of respondents (3/7) provided the correct diagnosis. The above-mentioned findings are all shown in Figure 3. Overall, these findings support the utility of our custom WSI pipeline for translational research activities.

**Figure 3:**
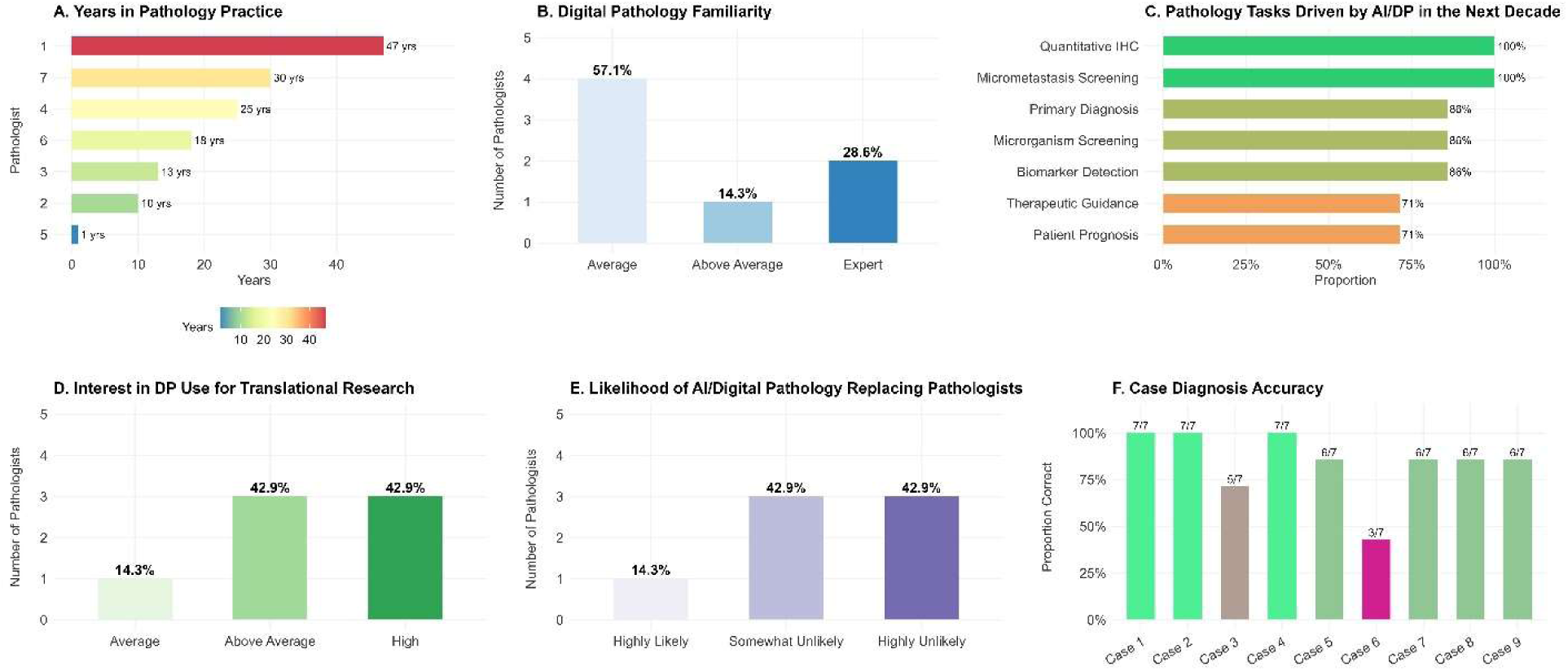
Pathologist survey outcomes A. Pathologist demographics B. Pathologist familiarity with digital pathology C. Pathologist perspectives on the common uses of AI/DP in pathology practice in the future. D. Pathologist responses for DP use in translational research. E. Likelihood of AI/DP replacing pathologists in the next decade F. Diagnostic accuracy of the nine cases used in this project across the pathologists

### D. Pathologist Questionnaire Data on the Custom WSI Platform

The pathologist user experience (UX) of our custom-built WSI platform was captured via a 23-question survey. The goals of the survey were divided based on the UX in two stages -i. A pre-use evaluation of the WSI platform access and navigation and ii. A post-use evaluation of the WSI platform usability and utility.

Part I (WSI platform access and navigation) - In this section, we addressed issues related to platform access and ease of use, as shown in questions 1-5 (see Figure 4). We considered this an important issue to address since our group consisted of three senior pathologists with over 25 years of pathology sign-out experience, with likely comfort with the analog processes. However, our custom WSI platform access and navigation were rated as easy or very easy by a majority of participants (71%), despite providing only minimal instructions: a website link and log-in details in an email (no verbal or individual instructions were provided). Pathologist users were provided freedom to explore the OMERO user interface (UI) without any explicit instructions. Overall, a majority of the pathologists (85%) felt the OMERO WSI portal provided no difficulty in visualization (very easy/easy, via zoom/pan) or loading the cases. The majority of respondents reported not using OMERO’s native annotation tool (86%). No specific instructions for using this annotation tool were provided, requiring pathologists to explore its features independently. Overall, the majority (71%) felt access to the portal presented no difficulty, which was encouraging.

**Figure 4:**
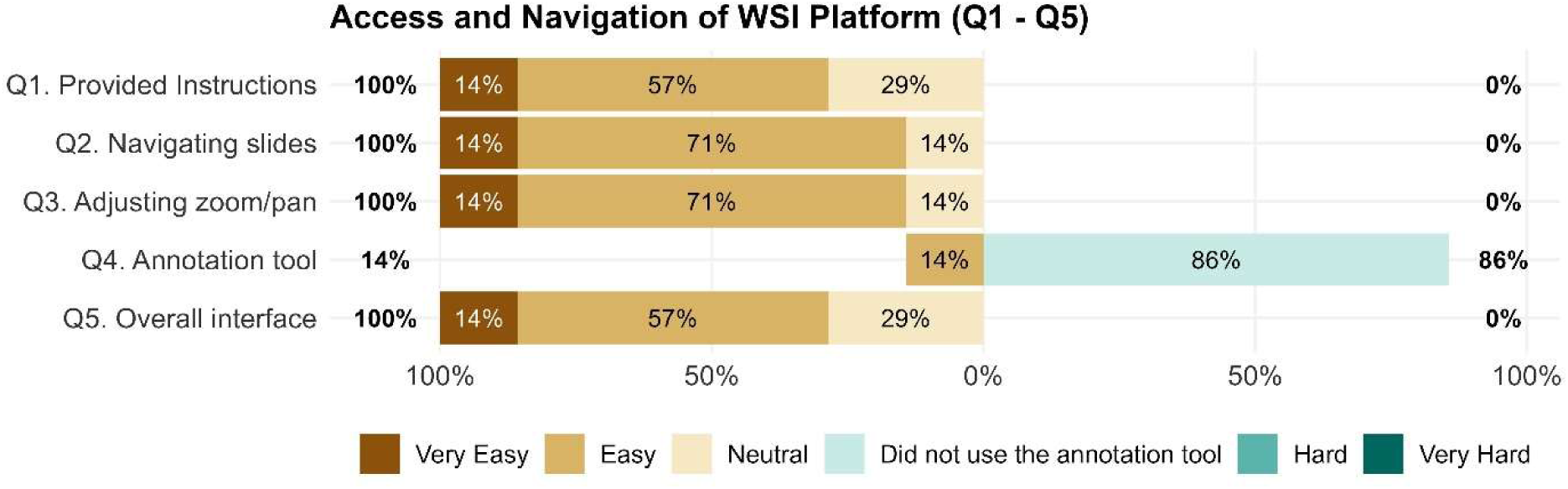
Pre-use user experience (UX) survey responses from the pathologists (Q1-Q5) focusing on ease of access/use to the WSI set up prior to and during slide reviews. Survey responses provided on a Likert scale (Very Easy- Easy-Neutral-Hard-Very Hard). Results shown as a % for each question response aggregated across the seven pathologists (n=7) who participated in the study.

Part II (WSI Usability and Utility) – In part II, we evaluated the post-use UX after working with our custom WSI setup. The pathologists were provided unlimited time (and access) to work with the OMERO WSI interface to get comfortable with the setup on their own schedule. In part II of the survey, we asked seventeen different survey questions focused on four topical areas – a. comparison to traditional microscopy workflow b. usability for translational research activity c. (potential) utility in a clinical pathology workflow and d. technical evaluation of the custom WSI pipeline In the initial section of the post-UX evaluation (Q6-Q10; comparison to traditional microscopy workflow), the pathologists were queried on their impressions of comparing the WSI setup to a traditional pathology microscopy workflow, which all of the pathologists were familiar with and used routinely. The majority of the pathologists did not feel that the setup either provided time savings (85%) nor was the image quality of the pipeline comparable to that of a traditional microscope (72%. The image quality issue may be due in part to insufficient optimization of the images used for the survey. We have made changes since then, resulting in higher-quality WSIs that will be used in the future (see Figure 1C). The key instance where the pathologists agreed was the utility of the WSI workflow in its potential for collaborative interactions (Q9), where a majority (86%) perceived value. The WSI workflow was perceived to be useful for multi-case reviews by equal numbers (Q8; 57% vs 43%). Lastly, all pathologists (Q10; 100%) felt more comfortable with traditional microscope-based workflow, illustrating a key shortcoming of building custom WSI pipelines, which likely lack the quality features of an advanced, polished commercial setup (at a high price point).

In the next part (Q11-Q15; usability for translational research), we queried the WSI set up usability for translational research activities, which was judged relatively favorably by the pathologists. The custom WSI set up was reviewed as being useful for – AI algorithm training (Q12; 86%), preferred platform for image-research studies (Q14; 71%), and translational research activities (Q15; 100%). A majority of pathologists did not use the OMERO annotation tool (Q11; 71%), while 29% who did use it found it useful. A majority of pathologists were neutral (Q13; 71%) about the idea that a DP workflow can increase communication between pathologists and the clinical team. Overall, we perceived a favorable outcome of the utility of custom WSI workflows such as ours in a low-risk setting, such as translational pathology research activities. Next, we surveyed the utility of our custom WSI workflow for clinical purposes (Q16-Q18; clinical pathology workflow utility). A majority (Q16; 57%) felt it could be used as a training ground for real-life clinical DP workflow implementation. Also, the majority (Q17; 71%) felt the custom WSI workflow could support clinical diagnostic workflow. However, the pathologists felt that the custom WSI setup was not quite ready for clinical practice with only minimal disruptions (Q18;71%).

In the last part (Q19-Q23; technical evaluation of the WSI pipeline), we sought to obtain feedback on the technical elements of the WSI workflow. In particular, we wanted to assess the value of utilizing open-source components to build such a WSI workflow. While the low cost of entry is indeed an attractive proposition for building such workflows, if the technical elements are unusable, the likelihood of adoption is low. Neither the WSI loading speed (Q19) nor the software interface (Q20) were perceived to be barriers by all pathologists. Since the images used in this project were primarily bone marrow samples, review of the fine histological details was considered to be of importance. A majority of the pathologists (Q21; 71%) perceived the WSI to be of concern at a high magnification in terms of the fine details of the image. This may be a function of the choice for the scanning that was adopted rather than the WSI quality itself (20X, 0.75 NA). The overall perception of the image quality of the WSI workflow was perceived to be poor by a slight majority compared to a traditional microscopy workflow (Q22; 57%). This is no surprise since an analog microscopy workflow has many more real-time microscope control adjustments to fine-tune the image compared to a WSI workflow. Lastly, a majority of the pathologists felt they could benefit from more training on the system (Q23; 71%) as opposed to the blind introduction to a custom WSI workflow that this project implemented. Survey results from Q6-Q23 are shown in Figure 5. Our overall impression of the pathologist feedback is one of measured optimism in light of the use of open-source, low-cost structure of this WSI workflow.

**Figure 5:**
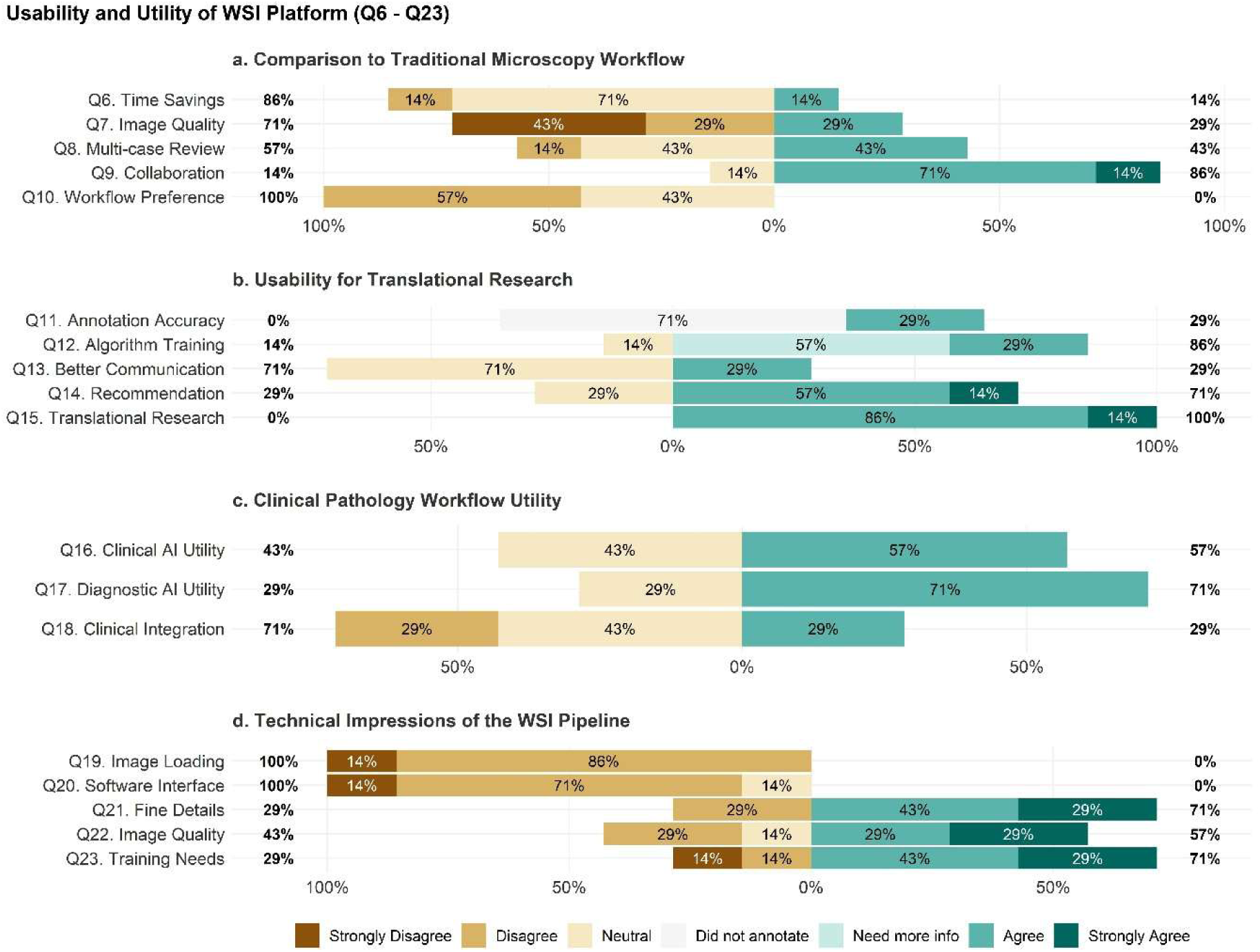
Post-use UX survey responses from the pathologists (Q6-Q23) surveying impressions of the custom WSI pipeline. Responses are divided into four subsections: a. Comparison to traditional pathology workflows (Q6-Q10), b. Usability for translational research (Q11-Q15), c. Clinical pathology workflow utility (Q16-Q18), and d. Technical evaluation of the WSI pipeline (Q19-Q23). UX survey responses provided on a Likert scale (Strongly Disagree- Disagree-Neutral-Need to know more-Agree-Strongly Agree). Results shown as a % for each question response aggregated across the seven pathologists (n=7) who participated in the study.

### IV. Discussion and Conclusions

Digitization of patient data is rapidly increasing across pathology, particularly in the context of anatomic pathology-driven histological slide processing and imaging^1,12,32,40^. Simultaneously, the concept of AI-enabled diagnostic automation and AI-assisted diagnosis is gaining favor and expanding in scope rapidly. These are changes happening right now in the landscape of pathology and healthcare that physicians and pathologists must prepare for. To ignore these winds of change would be to risk redundancy in the decades to come^1^. The convergence of AI automation and WSI foreshadows significant reimagining and restructuring of pathology practice over the next decade^8,41^. Nevertheless, there are many key challenges in the implementation of DP, such as the steep costs associated with scanner hardware purchases, trained personnel recruitment and implementation costs, recurring software purchase and maintenance costs, and the long-term management of digital data to ensure a robust and viable implementation of DP workflows in a clinical setting ^3,33^. All of these issues associated with DP workflows are non-trivial challenges in an era of constant cost-cutting measures and reduced medical reimbursements^32,42^. While the monetary gaps to establish DP workflows are real, the pressure faced by pathology practices to modernize and enable cutting-edge patient care is equally real. Pathologists (and departments) need to envision creative solutions to keep up with the latest approaches while maintaining reasonable budgets.

In the current study, we have demonstrated the technical approach, implementation, and evaluation of one such solution for digital pathology workflow implementation. We have established a low-cost, manual end-to-end WSI DP pipeline that may be suitable for small-scale laboratories and, more optimally, for translational pathology research initiatives. The goal of implementing this DP workflow setup is not to replace high-quality slide scanners. Instead, we sought to evaluate whether it could lower the technical and financial barriers for laboratories seeking to test DP workflows and protocols for the digitization of histologic slides. Our expert pathologists provided feedback that, while the setup in its current state would not be suitable for clinical applications, it would be highly appropriate for translational research activities. Over the course of this project, we prioritized open-source software at every step to increase its modular flexibility, accommodate individual laboratory preferences, and keep the total costs low. Although the current iteration of the WSI pipeline was judged by expert pathologists as having limited diagnostic utility, we expect this initial iteration can be significantly improved upon, to increase the quality of the WSIs and the user experience with additional technical optimization. Additionally, we sought to demonstrate the educational value of assembling the technical elements involved in a DP workflow that pathology trainees can learn from. We feel we were reasonably successful in achieving these modest goals of the project.

Over the course of the project, we identified multiple shortcomings of the custom-built DP pipeline. Our initial goals for the project included potential use of our DP workflow for clinical evaluative purposes (in addition to educational and translational research objectives). Diagnostic concordance between WSI based reviews and the gold standard of light microscopy in the recent literature has been reported to range from 90 to 95% ^9–11^. Our platform evaluated a small sample (n = 9) of bone marrow trephine biopsies with an impressive diagnostic concordance of 84% which was surprising to us. Additionally, routine diagnosis of MDS diagnostic entity is difficult by trephine biopsy histomorphology alone (as attempted in this project) and relies on the integration of clinical history, laboratory blood counts, peripheral smear review, aspirate smear, clot section, immunohistochemistry, cytogenetic, and molecular data ^43^. Based on this, it is unsurprising that our cohort of expert pathologists felt our WSI pipeline is unsuitable for routine clinical evaluative activities. The WSI issues are also further confounded by the relatively small region of interest (ROI) that was imaged, which was limited to 3 × 3 mm location. Furthermore, the pathologist survey results indicated poor cytologic detail of the scanned images which is key for hematological case evaluations which are often conducted at a 100X magnification. Although all of the slide images used in this study were captured using the same standard operating procedure (SOP), we identified slight variability between cases due to the manual acquisition procedures.

Further limitations of this study include the platform evaluation performed using a small sample (n = 7) of academic expert pathologists, many of whom are quite familiar with digital pathology technologies. And yet, despite being provided with limited instructions for accessing OMERO, the image management system, all of pathologists in the study were able to intuitively evaluate each case without additional queries or help. Another key limitation identified in this study is the relatively poor image quality of scanned slides, as judged by our expert pathologist panel. We feel this is primarily due to the use of a single 10× objective (0.4 NA) for image capture and WSI creation in this study. Additionally, pathologists are likely to have been exposed to higher-quality commercial WSIs than those used in this study, driving a perception of lower quality in this DP workflow. However, we feel these are technical challenges that can be overcome with further instrument optimization, which will be the focus of our future studies on this platform. The technical limitations precluding clinical use (as determined by our expert pathologists) are indeed issues we had expected from our non-automated, open-source DP imaging workflow. The pathologist feedback will serve as a baseline for future enhancements to improve the DP pipeline. We feel a majority of these technical shortcomings can be overcome with process, hardware, and software modifications.

An open-source infrastructure offers distinct advantages over closed, commercial, proprietary software. Unlike commercial end-to-end systems, modular open-source workflows enable targeted, community driven upgrades without needing to rely on expensive, vendor driven platform upgrades. Additionally, modular IT architectures provide greater flexibility for customers in terms of computational specifications, microscope-camera hardware options, and the use of community supported open-source management software. By having a greater say in the DP pipeline customization, end-users can drive WSI specification as needed by the end-user requirements rather than the vendor determined lock-in configuration. Separating the WSI hardware acquisition and WSI management systems components increases the flexibility and usability of WSI DP workflows without unnecessary feature bloat. Adopting open-source software workflows also eliminates the need for expensive, recurring maintenance contracts found in proprietary DP platforms, since the community can drive the key feature implementations as needed. While proprietary platforms do indeed offer distinct advantages (e.g., out of the box implementation and usability), open-source platforms are perhaps more robust for flexibility (at a cheaper price point) and can drive the competition for more economical options for widespread DP adoption. Our decision to develop an open-source DP workflow was validated by our survey pathologist experts, who judged our platform favorably for purposes of remote image review and translational pathology research collaborations. For smaller institutions with limited financial resources, an open-source platform like the one described in this paper could potentially lower the barriers to entry to DP and provide greater opportunities for experimentation with the imaging technologies to tailor to a diverse range of translational research needs, including multi-institutional collaborations and AI-enabled DP workflows which would otherwise be impossible due to the primary need for a commercial DP set up^1,44,45^.

Beyond its translational research applications, this open-source DP platform serves as an educational training ground for developing technical literacy, skills and experience in DP use^5,8^. As with the expectation that trainees must know how to gross a specimen and what occurs during routine histopathology processing, a technical understanding of the process of WSI creation (and its pitfalls) may soon become standard in pathology training. The American Board of Pathology has now fully transitioned to virtual microscopy, replacing exam questions based on glass slides. This change requires pathology trainees to have baseline competency with WSI handling in real-time. Our open-source DP pipeline offers ample educational opportunities to gain hands-on experience in WSI acquisition, processing, and subsequent handling. These kinds of educational exposures help trainees understand the importance of WSI technical concepts such as digital image file formats, image stitching algorithms, and components involved in server-based WSI management systems. Not only will these experiences prepare trainees for hybrid workflows, but they will also provide invaluable insights for those expected to manage laboratories with DP pipelines in the future. Such image-based competencies are becoming increasingly relevant as digitization becomes more widespread, and our open-source DP workflow model provides training programs with an opportunity to expose trainees without substantial capital investments.

The next steps to improve this open-source DP workflow involve addressing the major shortcomings identified in this project. These include - Evaluate the impact of WSI collections with higher-power, higher NA microscope objectives. The current iteration used an Olympus UPlanSApo 10×/0.40 objective, and our pathologist surveys indicated that image quality was perceived to be inferior to that of a traditional microscope workflow (Fig. 5). We can improve our DP pipeline through the use of higher magnification wide-field objectives, such as 20× and 40×, to increase the microscopic detail from the histological slides. The obvious trade-off will be a steep increase in the number of images needed to stitch with each higher objective to create a WSI, and therefore, larger file sizes. However, this could be overcome through the use of a more automated software pipeline for the image stitching process to create the WSIs. In the current study, we scanned a predefined 3 × 3 mm area to evaluate for features of MDS on trephine biopsies. Increasing the area of evaluation may provide greater histologic value for pathologist assessments than better nuclear and cytologic detail (at higher magnifications). Of course, a combination of higher-resolution objective images and increased surface area would provide the maximal utility. Identifying the best combinations (with pathologist review and feedback) is a critical first necessary step to systematically understand the bottlenecks associated with WSI file sizes and long-term server storage issues.

The current study solely evaluated bone marrow biopsies. However, many other tissue types are likely to have unique imaging requirements and remain to be investigated in detail. Differences in WSI outcomes as a function of tissue types, histology slide preparations, and scanning methodologies (e.g., magnification, real-time focusing strategies, and ROI selections) remains to be understood in detail. Detailed, systematic pathologist-feedback driven assessments are necessary to standardize the optical WSI/DP workflows in the future^42,44,46,47^. Understanding how these technical and software factors influence the final WSI quality and downstream diagnostic concordance would provide valuable insights into standard, optimal DP workflows suitable for clinical and translational pathology applications. Creating standardized WSI evaluation processes for a more diverse range of pathologists across various institutional settings and backgrounds may provide greater clarity on the expectations for digital microscopy, enabling rigorous quality control necessary for the universal adoption of DP workflows in pathology laboratories worldwide^42,44,46,47^.

Owing to the fully manual part of the microscope slide scanning and stitching parts of this DP workflow, the current setup is not optimized for speed of data acquisition. This part of the DP workflow can be automated (and sped up) through the use of motorized microscope stages. Computer controlled, motorized stages provide a more precise mechanism for moving glass slides along the X and Y axes, and in some hardware, even the Z axis. Compared to manual stages, a motorized stage scanning option is more accurate and reproducible than manual human control. These stages can be programmed to perform predefined, predictable scanning patterns with minimal user intervention. However, this adds an extra element of hardware and software expertise necessary that may not be readily available in a clinical laboratory. The aim of this study was to develop and assess cost-effective, manually operated WSI pipelines tailored for small-scale laboratories or research groups. Thus, the added and high costs of the motorized microscope stage option must be weighed against the need for a user-friendly DP platform option.

In summary, we have established an end-to-end, low-cost digital pathology workflow using readily available hardware components in a clinical pathology laboratory in conjunction with open-source image management software (i.e., OMERO). This feasibility study demonstrates the utility of such a DP platform for purposes of translational pathology research and educational activities in a routine clinical setting. Future work involves improving the quality of the DP pipeline through refinement of the optical microscopy hardware and custom software to improve the quality, speed, and usability of this DP workflow.

## Data Availability

All data produced in the present study are available upon reasonable request to the authors

## Authors’ contributions

Joseph Stenberg - Writing – original and revised draft, editing, data collection, data visualization, and editing. Aparna Gullapalli - software management, data visualization and editing. Kathryn Foucar – survey responses, writing – review and feedback. Daniel Babu – survey responses, writing – review and feedback. Jordan Redemann – survey responses, writing – review and feedback. Nancy Joste – survey responses, writing – review and feedback. Charles Foucar – writing – review and feedback. Dita Gratzinger – survey responses, writing – review and feedback. Tracy George – survey responses, writing – review and feedback. Robert Ohgami – survey responses, writing – review and feedback. Rama R. Gullapalli - writing – original and revised draft, data collection, data visualization, software management, editing, project supervision, funding acquisition, and idea conceptualization.

## Funding Acknowledgments

RRG was supported by a grant from the UNM Center for Metals in Biology and Medicine (CMBM) through NIH NIGMS grant P20 GM130422.

---

**Supplemental Figure 1:**
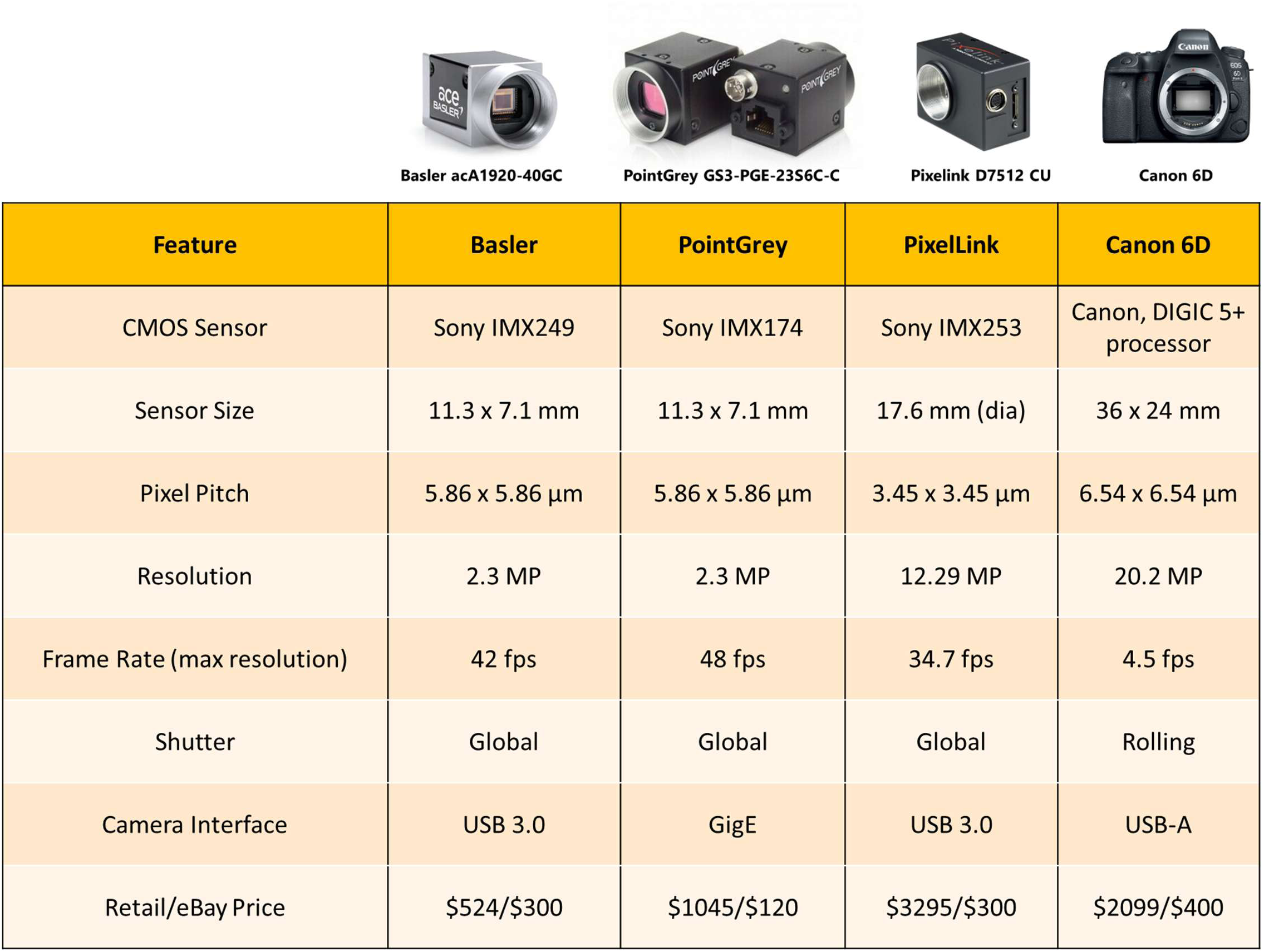
The specimen review process for inclusion of bone marrow biopsy cases used in the current custom WSI pipeline project.

**Supplemental Figure 2:**
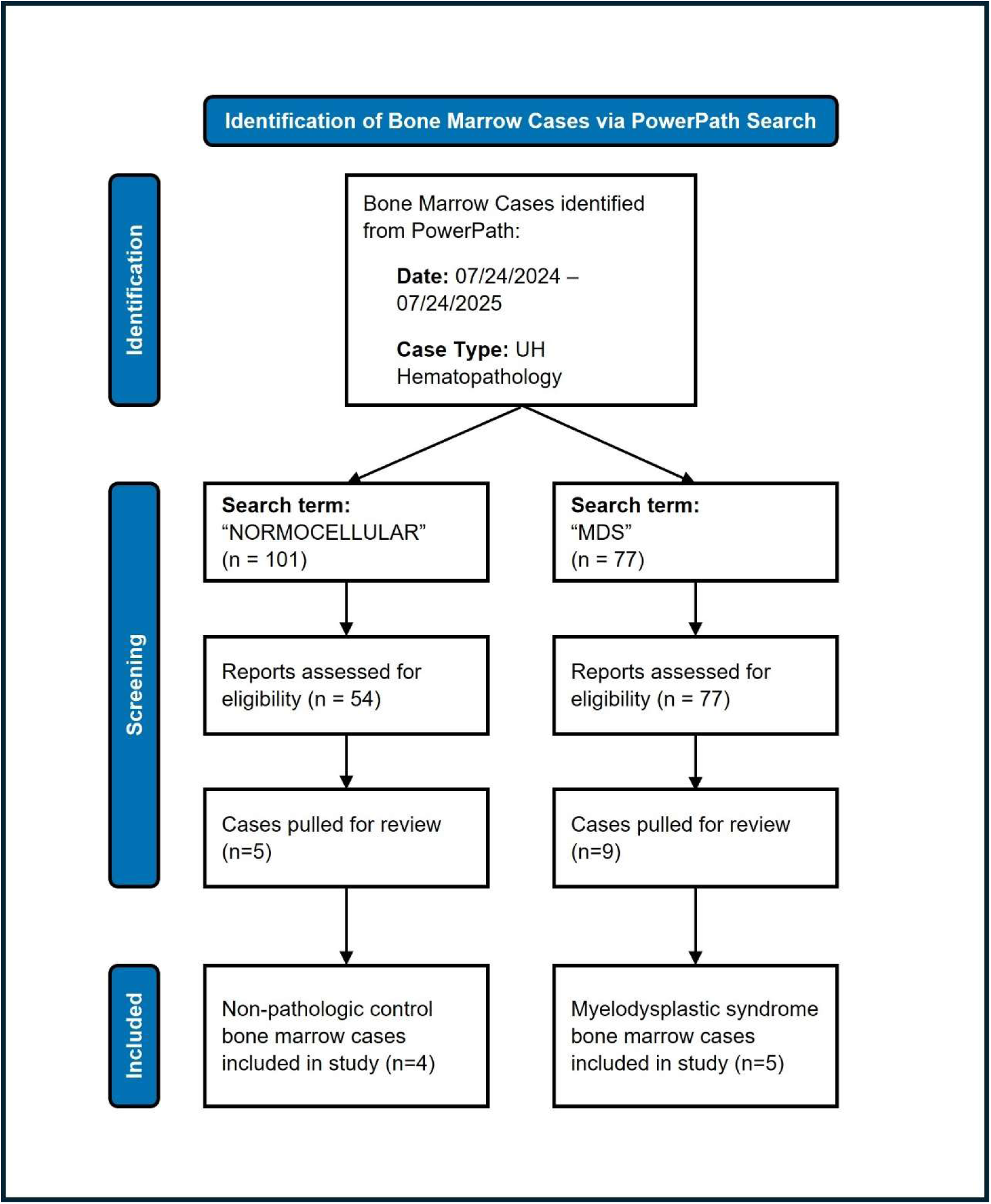
Technical details and pricing of the various cameras surveyed for use in the current project. The Canon 6D camera was chosen for the ease of use, full-frame sensor, and quality of images collected

